# Left ventricular mass-to-strain ratio to predict treatment response and prognosis in hypertensive heart disease

**DOI:** 10.1101/2025.02.25.25322843

**Authors:** In-Chang Hwang, Hyue Mee Kim, Jiesuck Park, Hong-Mi Choi, Yeonyee E. Yoon, Goo-Yeong Cho

**Author notes:** These two authors equally contributed to this work as co-corresponding authors (In-Chang Hwang, Hyue Mee Kim). **Conflict of Interest:** The authors declare no conflicts of interest. **Address for correspondence:** In-Chang Hwang, MD Associate Professor Department of Internal Medicine, Seoul National University College of Medicine Cardiovascular Center, Seoul National University Bundang Hospital 82 Gumi-ro-173-gil, Bundang, Seongnam, Gyeonggi, 13620, South Korea / *and* Hyue Mee Kim, MD Assistant Professor, Division of Cardiology, Department of Internal Medicine, Chung-Ang University Hospital, Chung-Ang University College of Medicine 102, Heukseok-ro, Dongjak-gu, Seoul, 06973, South Korea /.

## Abstract

**Background:** Typical hypertensive heart disease (HHD) phenotype is the left ventricular hypertrophy (LVH) and dysfunction, measurable via LV global longitudinal strain. We aimed to evaluate the LV mass-to-strain ratio (LV-MSR) as a marker of HHD treatment response and prognosis.

**Methods:** We retrospectively analyzed consecutive patients who underwent echocardiography at hypertension diagnosis and 6–18-monthly intervals in tertiary centers from 2006–2021. Association between LV-MSR and LV geometry changes was assessed using receiver operating characteristic (ROC) analysis and multivariable logistic regression. Time-dependent ROC and Cox regression were performed to evaluate LV-MSR prognostic value for cardiovascular death and heart failure hospitalization.

**Results:** Among 1,600 patients (mean age, 65.1 years; 61.1% male), 23.4% had concentric LVH, and 21.5% eccentric LVH at baseline. LV-MSR had the highest predictive accuracy for LV geometry changes (area-under-the-curve 0.786; 95% confidence interval [CI] 0.759– 0.813). LV-MSR showed significant association with new-onset LVH among the non-LVH group (adjusted odds ratio, 1.125, 95%CI 1.054–1.200, P<0.001) and persistent LVH among LVH group (adjusted hazard ratio [aHR] 1.133, 95%CI 1.087–1.180, P<0.001) at baseline. Higher LV-MSR was an independent prognosticator, whether as a continuous (aHR 1.032 per +1 g/m^2^/%, 95%CI 1.015–1.049, P<0.001) or categorical variable (aHR 2.257 for LV-MSR ≥6.52 g/m^2^/%, 95%CI 1.463–3.480, P<0.001). The associations persisted in subgroup analyses according to the presence of LVH baseline.

**Conclusions:** LV-MSR independently predicts LV geometry changes and clinical outcomes in HHD, serving as a superior prognostic marker compared to LV mass index or LV-GLS alone.

**Clinical Perspective:** *What is new?:*

- LV mass-to-strain ratio (LV-MSR) integrates LV geometry (LVH) and LV longitudinal function (LV-GLS), providing a more comprehensive assessment of hypertensive heart disease (HHD) than either parameter alone.

*What are the clinical implications?:*

- Higher LV-MSR predicts LVH progression, persistence, and worse clinical outcomes, making it a useful marker for risk stratification and treatment response assessment in hypertension.

*Research Perspective:*

- Further studies should validate LV-MSR across diverse populations, integrate it into standard echocardiographic protocols, and evaluate its role in clinical decision-making and antihypertensive treatment guidance to improve patient outcomes.

## Introduction

The hallmark phenotype of hypertensive heart disease (HHD) is an increased left ventricular (LV) mass, which results from a prolonged increase in the afterload applied to the LV myocardium.^1^ Additionally, the increased LV mass in HHD is often complicated by LV systolic or diastolic dysfunction.^2^ Considering that the fundamental goal of adaptive LV structural remodeling in HHD is to maintain appropriate cardiac output in compensation of the increased afterload, the patients with HHD often do not show significant abnormalities in conventional echocardiographic parameters.^3^ Instead, the LV global longitudinal strain (LV-GLS) has been suggested as a potential indicator for both overt and subclinical myocardial dysfunction in HHD.^4-6^ A meta-analysis of 4,276 individuals (2,187 patients with hypertension) reported that the LV ejection fraction (LV-EF) did not differ between normotensive and hypertensive patients; however, the LV-GLS was significantly impaired in the hypertensive group.^7^

Considering that LV myocardium undergoes hypertrophic remodeling to compensate for the increased afterload, while sacrificing the longitudinal contraction, the relationship between the degree of LV hypertrophy (LVH; i.e., LV mass) and degree of attenuation of myocardial longitudinal motion (i.e., LV-GLS) may potentially reflect better the dynamic status of LV remodeling following the increased afterload. Therefore, the LV mass-to-strain ratio (LV-MSR) holds promise as an indicator of the consequence of LV remodeling in patients with HHD. Several studies suggest the clinical usefulness of the LV-MSR in various myocardial conditions.^8,9^ However, no previous study has assessed the use of LV-MSR as an indicator of response to antihypertensive treatment, or as a prognostic marker, in patients with HHD. In this explorative analysis of echocardiographic findings in patients with HHD, we aimed to investigate whether the LV-MSR could be an ideal indicator for the progression or regression of LVH, and be used as a prognostic marker.

## Methods

### Study population

Participants comprised consecutive patients with hypertension who underwent an initial echocardiography during hypertension diagnosis at Seoul National University Bundang Hospital or Chung-Ang University Hospital (tertiary care centers in Korea) between 2006 and 2021, and completed a follow-up echocardiography during antihypertensive treatment at 6– 18-monthly intervals.^10-12^ Exclusion criteria were: (1) specific cardiomyopathies, such as dilated cardiomyopathy, hypertrophic cardiomyopathy, restrictive cardiomyopathy, ischemic cardiomyopathy, stress-induced cardiomyopathy, Fabry disease, and MELAS (mitochondrial encephalopathy, lactic acidosis, and stroke-like episodes); (2) end-stage renal disease; and (3) prior open-heart surgery. Among the 1,872 patients who were included after the above eligibility criteria, patients without available echocardiograms for LV-GLS measurement were further excluded. Finally, a total of 1,600 patients were included.

This study was conducted in accordance with the principles of the Declaration of Helsinki and approved by the institutional review board of the Clinical Research Institute at each hospital (Seoul National University Bundang Hospital: No. B-2206-762-102, Chung-Ang University Hospital: No. 2205-014-19419). Given the retrospective nature of the study and minimal expected risk to the participants, the requirement for informed consent was waived by the institutional review boards. The study protocol was registered with the Clinical Research Information Service of the Ministry of Health and Welfare of the Republic of Korea (No. KCT0008091).

### Echocardiography

All patients underwent conventional two-dimensional M-mode and color Doppler examinations using a commercially available echocardiographic device with a 2-2.5 MHz transducer. Echocardiographic parameters were measured according to the American Society of Echocardiography guidelines.^13^ LV end-diastolic dimension (LV-EDD), LV end-systolic dimension (LV-ESD), and LV septal and posterior wall thicknesses were estimated using M-mode or two-dimensional echocardiography. LV mass was measured using Devereux’s formula, and the LV mass index (LV-MI) was calculated by normalization to the body surface area. LVH was defined as LV-MI >115 g/m^2^ and >95 g/m^2^ in men and women, respectively.^14^ The relative wall thickness (RWT) was the ratio of twice the posterior wall thickness divided by the LV-EDD; values >0.42 indicated an increased RWT. To obtain the LV-EF, LV end-diastolic and LV end-systolic volumes were measured using Simpson’s biplane method in the apical two-chamber and four-chamber views. The left atrial (LA) volume index was calculated by dividing the LA volume by the body surface area. The right ventricular systolic pressure was calculated from the peak velocity of tricuspid regurgitation with right atrial pressure. LV-GLS was analyzed from apical 4-, 2-, and 3-chamber views using TomTec Image-Arena, and was expressed as the absolute value for ease in interpretation.^13^

### Outcomes

The LV geometry outcomes were the occurrences of LVH or persistent LVH, based on the follow-up echocardiogram among patients without and with LVH at baseline, respectively. The clinical outcome was a composite of cardiovascular death (CV death) and hospitalization for heart failure (HHF). HHF was defined as inpatient admission owing to worsening HF signs and symptoms. Data on the clinical outcome were obtained by reviewing individual hospital records, telephone contracts, and national mortality data.

### Statistical analysis

Continuous variables are presented as the mean ± standard deviation and categorical variables as frequencies. Student’s independent t-test and Kruskal–Wallis test were used to examine the clinical and echocardiographic characteristics. Logistic regression analysis was performed to identify predictors of new-onset LVH (among patients without LVH at baseline) and persistent LVH (among the patients with LVH at baseline). Univariable factors with P-values <0.100 were entered into the multivariable logistic regression analysis models using the stepwise backward selection method. The adjusted odds ratios (OR) for the new-onset LVH or persistent LVH were visualized by LV-MSR at baseline, using the cubic spline curves. Receiver-operating characteristics (ROC) curve analysis with DeLong test was used to assess the predictive value of baseline echocardiographic parameters for the presence of LVH at follow-up echocardiogram, and time-dependent ROC curve analysis with bootstrap-based method was used to assess the prognostic values of baseline echocardiographic parameters for the occurrence of clinical outcomes. Event-free survival analyses were performed using the Kaplan-Meier method and Cox proportional hazard modeling. Variables found to be significant in univariable analysis were entered into a multivariable Cox proportional-hazards regression model using the stepwise backward selection method. Mediation analysis was performed to assess the direct and indirect effects of LV-MSR on the clinical outcomes, using R package “mediation”. All analyses were performed using R programming software (version 4.3.3; R Foundation for Statistical Computing, Vienna, Austria), and P-values <0.05 were considered statistically significant.

## Results

### Clinical characteristics

The mean age of the total study population was 65.1 years (SD 13.1), 61.1% were male, and the prevalence of diabetes, dyslipidemia, chronic kidney disease, and coronary artery disease was 28.9%, 28.1%, 23.2%, and 28.1%, respectively (**Table 1**). Baseline characteristics were compared between those without (n=882) and with (n=714) LVH at baseline. The mean age and prevalence of diabetes, dyslipidemia, atrial fibrillation, and stroke were similar between the two groups. When compared to those without LVH at baseline, those with LVH at baseline had significantly lower male proportion (70.3% vs. 49.7%; P<0.001) and lower prevalence of dyslipidemia (30.7% vs. 24.8%; P=0.009) and coronary artery disease (41.6% vs. 33.7%; P=0.001), and higher prevalence of chronic kidney disease (19.0% vs. 28.4%; P<0.001). The blood pressure (BP) at baseline was significantly higher in patients with LVH compared to those without LVH (systolic BP [SBP], 148.6±20.2 vs. 158.3±27.1 mmHg, P<0.001; diastolic BP [DBP], 87.3±14.6 vs. 92.6±21.3 mmHg, P<0.001).

**Table 1.** Clinical characteristics.

| Variables | Total population<br>(n=1,600) | No LVH at baseline<br>(n=882) | LVH at baseline<br>(n=714) | P |
| --- | --- | --- | --- | --- |
| **Clinical Factors** |  |  |  |  |
| Age (years) | 65.1 ± 13.1 | 65.1 ± 12.3 | 65.1 ± 14.1 | 0.965 |
| Male sex | 977 (61.1%) | 620 (70.3%) | 357 (49.7%) | <0.001 |
| Diabetes mellitus | 462 (28.9%) | 247 (28.0%) | 215 (29.9%) | 0.406 |
| Dyslipidemia | 449 (28.1%) | 271 (30.7%) | 178 (24.8%) | 0.009 |
| Chronic kidney disease | 372 (23.2%) | 168 (19.0%) | 204 (28.4%) | <0.001 |
| Coronary artery disease | 609 (38.1%) | 367 (41.6%) | 242 (33.7%) | 0.001 |
| Atrial fibrillation | 222 (13.9%) | 116 (13.2%) | 106 (14.8%) | 0.345 |
| Stroke | 210 (13.1%) | 110 (12.5%) | 100 (13.9%) | 0.413 |
| **Anthropometric Factors** |  |  |  |  |
| Body mass index (kg/m<sup>2</sup>) | 25.2 ± 3.6 | 25.1 ± 3.5 | 25.3 ± 3.8 | 0.226 |
| SBP at baseline (mmHg) | 152.9 ± 24.0 | 148.6 ± 20.2 | 158.3 ± 27.1 | <0.001 |
| SBP at follow-up (mmHg) | 129.4 ± 16.1 | 127.8 ± 15.4 | 131.4 ± 16.8 | <0.001 |
| ΔSBP (mmHg) | -23.5 ± 24.4 | -20.8 ± 21.8 | -26.9 ± 26.9 | <0.001 |
| DBP at baseline (mmHg) | 89.7 ± 18.1 | 87.3 ± 14.6 | 92.6 ± 21.3 | <0.001 |
| DBP at follow-up (mmHg) | 82.3 ± 23.2 | 80.7 ± 16.2 | 84.3 ± 29.6 | 0.004 |
| ΔDBP (mmHg) | -7.7 ± 21.3 | -6.6 ± 18.8 | -9.1 ± 23.9 | 0.021 |
| Heart rate (bpm) | 71.8 ± 21.0 | 70.5 ± 21.5 | 73.5 ± 20.3 | 0.005 |
| **Laboratory Findings** |  |  |  |  |
| Hemoglobin (g/dL) | 13.4 ± 2.1 | 13.6 ± 1.9 | 13.1 ± 2.2 | <0.001 |
| Blood urea nitrogen (mg/dL) | 17.9 ± 8.9 | 16.9 ± 7.2 | 19.3 ± 10.5 | <0.001 |
| Serum creatinine (mg/dL) | 1.0 ± 0.7 | 0.9 ± 0.4 | 1.1 ± 0.9 | <0.001 |
| GFR (mL/min/1.73m<sup>2</sup>) | 79.5 ± 26.0 | 82.9 ± 24.3 | 75.2 ± 27.3 | <0.001 |
| Total cholesterol (mg/dL) | 168.5 ± 44.9 | 166.2 ± 44.5 | 171.2 ± 45.2 | 0.028 |
| Triglyceride (mg/dL) | 131.5 ± 88.0 | 134.2 ± 91.2 | 128.1 ± 83.9 | 0.180 |
| HDL cholesterol (mg/dL) | 46.5 ± 12.9 | 46.1 ± 12.6 | 47.1 ± 13.2 | 0.148 |
| LDL cholesterol (mg/dL) | 101.1 ± 35.8 | 100.1 ± 38.0 | 102.4 ± 33.0 | 0.202 |
| **Antihypertensive medication** |  |  |  |  |
| RAS blockers | 1071 (66.9%) | 558 (63.3%) | 513 (71.4%) | 0.001 |
| Beta blockers | 582 (36.4%) | 306 (34.7%) | 276 (38.4%) | 0.130 |
| DHP-CCB | 735 (45.9%) | 357 (40.5%) | 378 (52.6%) | <0.001 |
| NDHP-CCB | 96 (6.0%) | 59 (6.7%) | 37 (5.2%) | 0.206 |
| Thiazide | 261 (16.3%) | 102 (11.6%) | 159 (22.1%) | <0.001 |
| Alpha blockers | 28 (1.8%) | 9 (1.0%) | 19 (2.6%) | 0.020 |
| MRA | 122 (7.6%) | 28 (3.2%) | 94 (13.1%) | <0.001 |
| Loop diuretics | 210 (13.1%) | 65 (7.4%) | 145 (20.2%) | <0.001 |

Values are given as the mean ± standard deviation or number with percentage (%).
Abbreviations: DBP, diastolic blood pressure; DHP-CCB, dihydropyridine calcium channel blocker; GFR, glomerular filtration rate; HDL, high-density lipoprotein; LDL, low-density lipoprotein; LVH, left ventricular hypertrophy; MRA, mineralocorticoid antagonist; NDHP-CCB, non-dihydropyridine calcium channel blocker; RAS, renin-angiotensin system; SBP, systolic blood pressure

Echocardiographic parameters at baseline and follow-up are summarized in **Table 2**. The mean LV-MI was 109.8±33.7 g/m^2^ (patients without vs. with LVH at baseline, 88.7±14.1 g/m^2^ vs. 135.7±32.8 g/m^2^; P<0.001), while the mean LV-GLS was 15.3±5.0% (patients without vs. with LVH at baseline, 16.4±4.6% vs. 14.1±5.2%, P<0.001). The mean LV-MI at follow-up did not significantly change in patients without LVH at baseline; however, in patients with LVH at baseline, the follow-up LV-MI was markedly decreased at follow-up (LV-MI at follow-up, 97.7±26.4 vs. 123.3±35.5 g/m^2^; P<0.001; △LV-MI, 9.0±25.0 vs. - 12.5±38.7 g/m^2^; P<0.001). Among the 882 patients without LVH at baseline, 220 (24.9%) developed LVH at follow-up (including 103 [11.7%] showed concentric LVH, and 117 [13.3%] eccentric LVH). Among the 714 patients with LVH at baseline, 237 patients (33.2%) showed regression of LVH at follow-up (including 147 [20.5%] showed normal geometry, and 90 [12.5%] concentric remodeling).

**Table 2.**
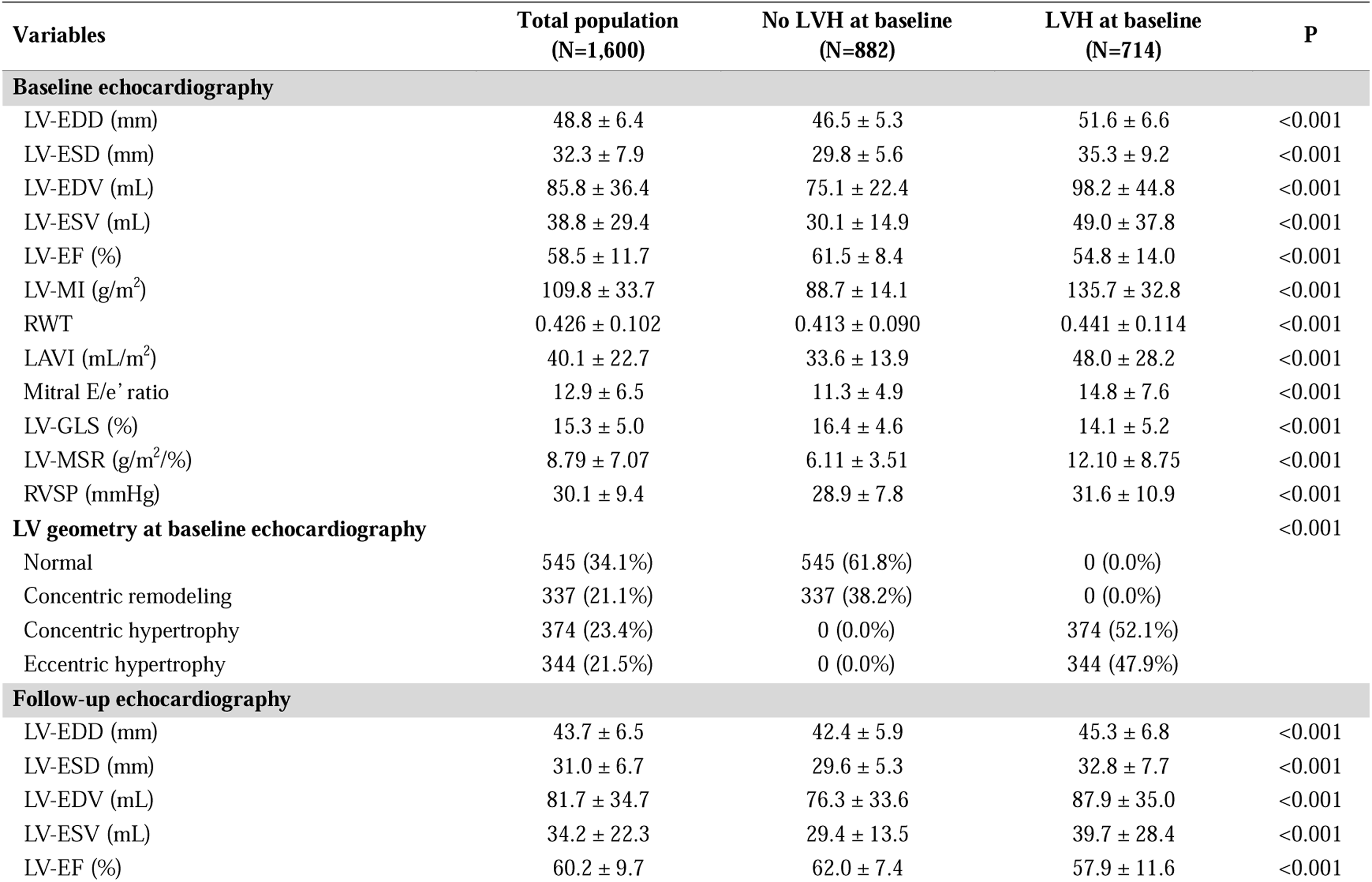
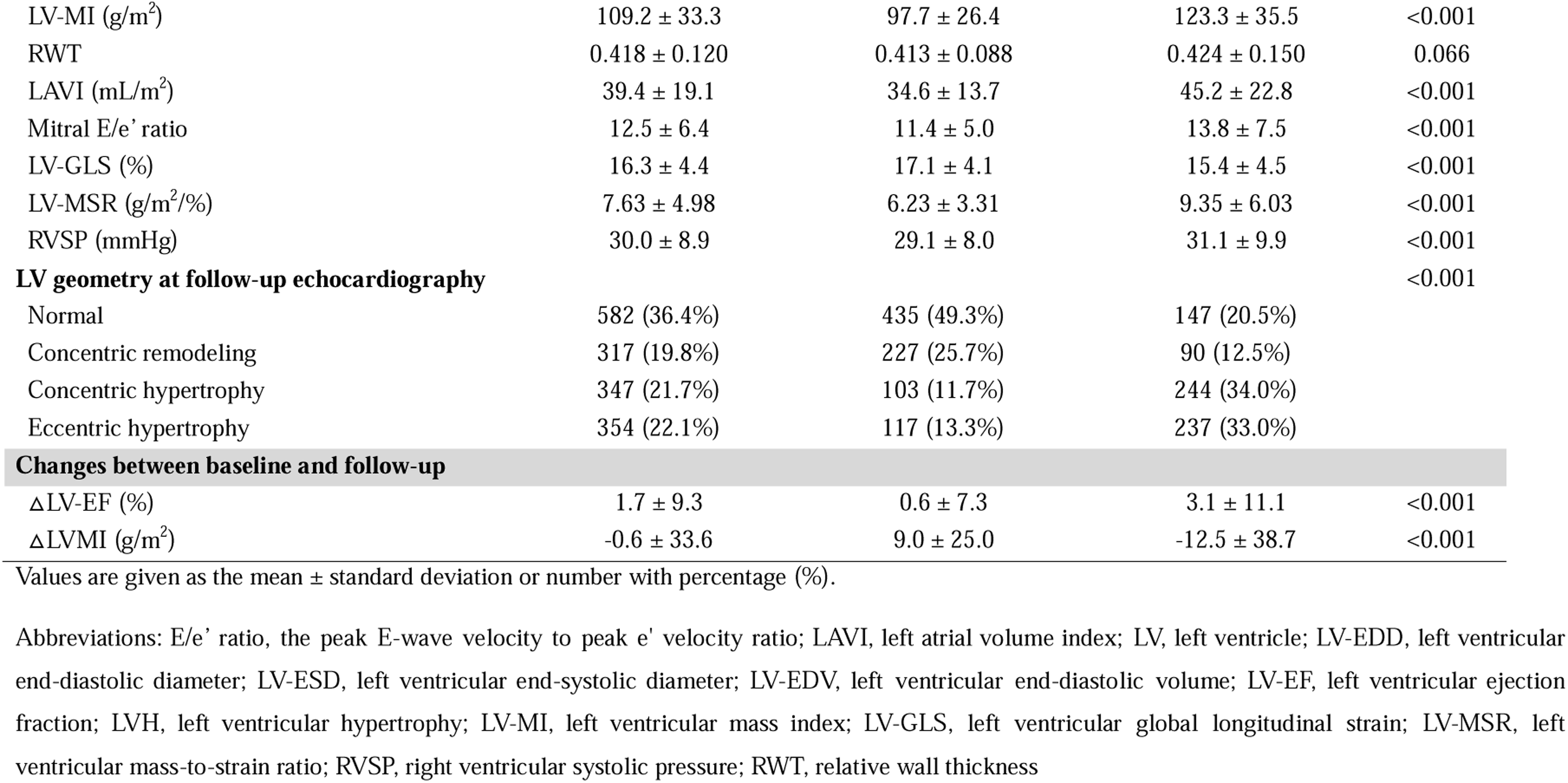
Echocardiographic parameters at baseline and follow-up assessment.

| Variables | Total population<br>(N=1,600) | No LVH at baseline<br>(N=882) | LVH at baseline<br>(N=714) | P |
| --- | --- | --- | --- | --- |
| **Baseline echocardiography** |  |  |  |  |
| LV-EDD (mm) | 48.8 ± 6.4 | 46.5 ± 5.3 | 51.6 ± 6.6 | <0.001 |
| LV-ESD (mm) | 32.3 ± 7.9 | 29.8 ± 5.6 | 35.3 ± 9.2 | <0.001 |
| LV-EDV (mL) | 85.8 ± 36.4 | 75.1 ± 22.4 | 98.2 ± 44.8 | <0.001 |
| LV-ESV (mL) | 38.8 ± 29.4 | 30.1 ± 14.9 | 49.0 ± 37.8 | <0.001 |
| LV-EF (%) | 58.5 ± 11.7 | 61.5 ± 8.4 | 54.8 ± 14.0 | <0.001 |
| LV-MI (g/m<sup>2</sup>) | 109.8 ± 33.7 | 88.7 ± 14.1 | 135.7 ± 32.8 | <0.001 |
| RWT | 0.426 ± 0.102 | 0.413 ± 0.090 | 0.441 ± 0.114 | <0.001 |
| LAVI (mL/m<sup>2</sup>) | 40.1 ± 22.7 | 33.6 ± 13.9 | 48.0 ± 28.2 | <0.001 |
| Mitral E/e' ratio | 12.9 ± 6.5 | 11.3 ± 4.9 | 14.8 ± 7.6 | <0.001 |
| LV-GLS (%) | 15.3 ± 5.0 | 16.4 ± 4.6 | 14.1 ± 5.2 | <0.001 |
| LV-MSR (g/m<sup>2</sup>/%) | 8.79 ± 7.07 | 6.11 ± 3.51 | 12.10 ± 8.75 | <0.001 |
| RVSP (mmHg) | 30.1 ± 9.4 | 28.9 ± 7.8 | 31.6 ± 10.9 | <0.001 |
| **LV geometry at baseline echocardiography** |  |  |  | <0.001 |
| Normal | 545 (34.1%) | 545 (61.8%) | 0 (0.0%) |  |
| Concentric remodeling | 337 (21.1%) | 337 (38.2%) | 0 (0.0%) |  |
| Concentric hypertrophy | 374 (23.4%) | 0 (0.0%) | 374 (52.1%) |  |
| Eccentric hypertrophy | 344 (21.5%) | 0 (0.0%) | 344 (47.9%) |  |
| **Follow-up echocardiography** |  |  |  |  |
| LV-EDD (mm) | 43.7 ± 6.5 | 42.4 ± 5.9 | 45.3 ± 6.8 | <0.001 |
| LV-ESD (mm) | 31.0 ± 6.7 | 29.6 ± 5.3 | 32.8 ± 7.7 | <0.001 |
| LV-EDV (mL) | 81.7 ± 34.7 | 76.3 ± 33.6 | 87.9 ± 35.0 | <0.001 |
| LV-ESV (mL) | 34.2 ± 22.3 | 29.4 ± 13.5 | 39.7 ± 28.4 | <0.001 |
| LV-EF (%) | 60.2 ± 9.7 | 62.0 ± 7.4 | 57.9 ± 11.6 | <0.001 |
| LV-MI (g/m<sup>2</sup>) | 109.2 ± 33.3 | 97.7 ± 26.4 | 123.3 ± 35.5 | <0.001 |
| RWT | 0.418 ± 0.120 | 0.413 ± 0.088 | 0.424 ± 0.150 | 0.066 |
| LAVI (mL/m<sup>2</sup>) | 39.4 ± 19.1 | 34.6 ± 13.7 | 45.2 ± 22.8 | <0.001 |
| Mitral E/e' ratio | 12.5 ± 6.4 | 11.4 ± 5.0 | 13.8 ± 7.5 | <0.001 |
| LV-GLS (%) | 16.3 ± 4.4 | 17.1 ± 4.1 | 15.4 ± 4.5 | <0.001 |
| LV-MSR (g/m<sup>2</sup>%) | 7.63 ± 4.98 | 6.23 ± 3.31 | 9.35 ± 6.03 | <0.001 |
| RVSP (mmHg) | 30.0 ± 8.9 | 29.1 ± 8.0 | 31.1 ± 9.9 | <0.001 |
| **LV geometry at follow-up echocardiography** |  |  |  | <0.001 |
| Normal | 582 (36.4%) | 435 (49.3%) | 147 (20.5%) |  |
| Concentric remodeling | 317 (19.8%) | 227 (25.7%) | 90 (12.5%) |  |
| Concentric hypertrophy | 347 (21.7%) | 103 (11.7%) | 244 (34.0%) |  |
| Eccentric hypertrophy | 354 (22.1%) | 117 (13.3%) | 237 (33.0%) |  |
| **Changes between baseline and follow-up** |  |  |  |  |
| ΔLV-EF (%) | 1.7 ± 9.3 | 0.6 ± 7.3 | 3.1 ± 11.1 | <0.001 |
| ΔLVMI (g/m<sup>2</sup>) | -0.6 ± 33.6 | 9.0 ± 25.0 | -12.5 ± 38.7 | <0.001 |

Values are given as the mean ± standard deviation or number with percentage (%).
Abbreviations: E/e' ratio, the peak E-wave velocity to peak e' velocity ratio; LAVI, left atrial volume index; LV, left ventricle; LV-EDD, left ventricular end-diastolic diameter; LV-ESD, left ventricular end-systolic diameter; LV-EDV, left ventricular end-diastolic volume; LV-EF, left ventricular ejection fraction; LVH, left ventricular hypertrophy; LV-MI, left ventricular mass index; LV-GLS, left ventricular global longitudinal strain; LV-MSR, left ventricular mass-to-strain ratio; RVSP, right ventricular systolic pressure; RWT, relative wall thickness

### LV mass-to-strain ratio to determine the changes in LV geometry

We performed ROC curve analysis to determine the optimal echocardiographic predictor of the changes in LV geometry. As shown in **Figure 1**, the LV-MSR at baseline had the highest area under the curve (AUC 0.786, 95% CI 0.759–0.813; P<0.001), followed by LV-MI (AUC 0.735, 95% CI 0.704–0.766; P<0.001), E/e’ ratio (AUC 0.668, 95% CI 0.634– 0.703; P<0.001), and LV-GLS (AUC 0.625, 95% CI 0.589–0.611; P<0.001). The AUC for the presence of LVH at follow-up echocardiogram was significantly higher with the LV-MSR, compared to LV-GLS and LV-MI.

**Figure 1.**
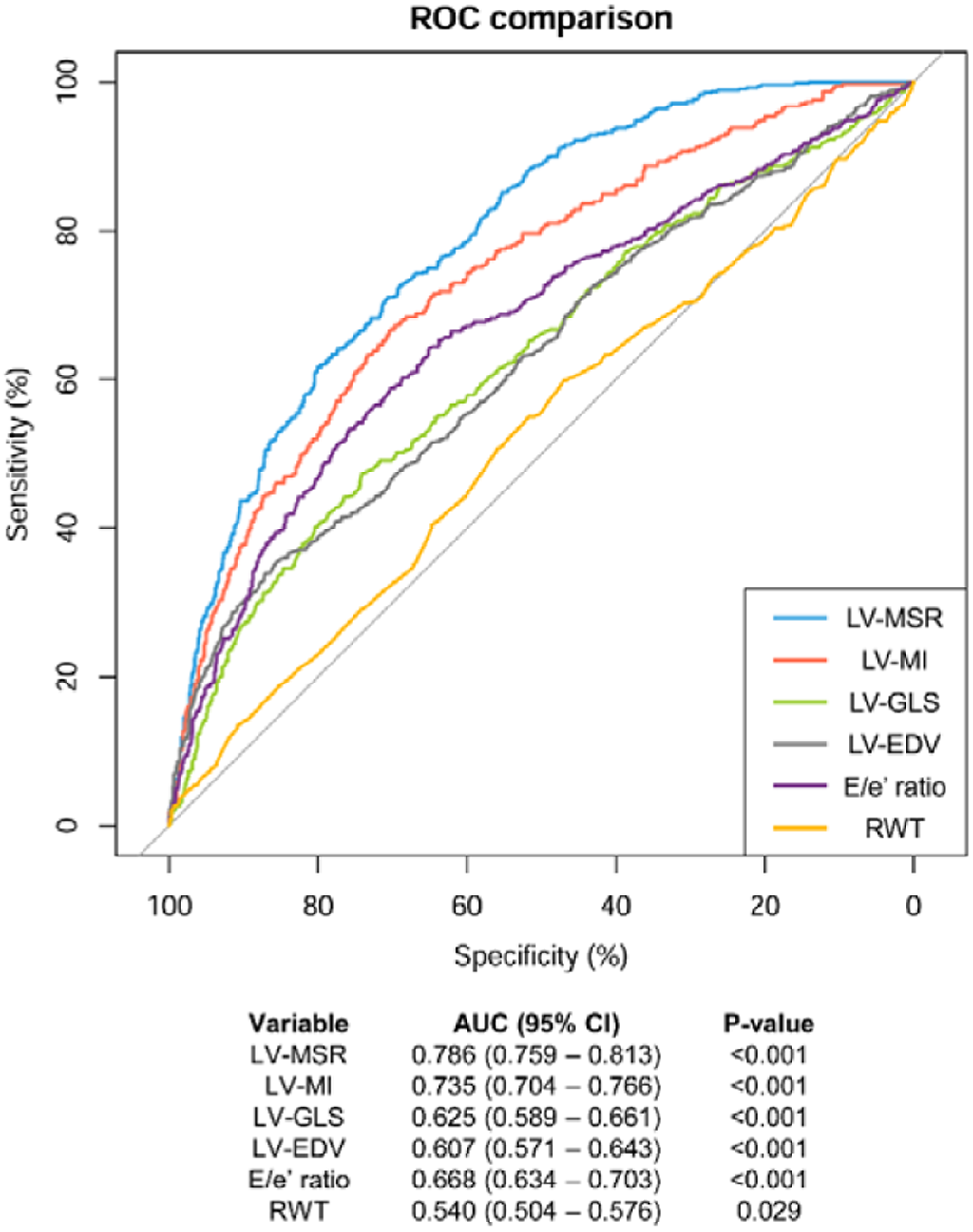
ROC curve analysis of echocardiographic parameters to predict new-onset or persistent LVH. Abbreviations: ROC, receiver operating characteristic; LV-MSR, left ventricular mass-to-strain ratio; LV-MI, left ventricular mass index; LV-GLS, left ventricular global longitudinal strain; LV-EDV, left ventricular end-diastolic volume; RWT, relative wall thickness; AUC, area under the curve; CI, confidence interval.

Multiple linear regression analysis was performed to assess the association between baseline echocardiographic parameters and the degree of LV-MI reduction (△LV-MI) between the baseline and follow-up echocardiograms (**Supplemental Table S1**). Higher BMI at baseline was associated with less reduction in LV-MI, whereas higher SBP at baseline was associated with larger reduction in LV-MI. Among the echocardiographic parameters at baseline, larger LV-EDV (adjusted coefficient β -0.204, 95% CI -0.262 to -0.145; P<0.001), higher RWT (adjusted coefficient β -60.589, 95% CI -77.243 to -43.935; P<0.001), and higher LV-MSR (adjusted coefficient β -0.602, 95% CI -0.875 to -0.330; P<0.001) were associated with a larger reduction in LV-MI.

### Subgroup analyses according to the presence of LVH at baseline

The association between LV-MSR at baseline and changes in LV geometry was further assessed in subgroups divided according to the presence of LVH at baseline (**Table 3**). Among patients without LVH at baseline, higher LV-MSR was significantly associated with new-onset LVH at follow-up (adjusted OR 1.125, 95% CI 1.054–1.200; P<0.001), as well as higher RVSP. Among those with LVH at baseline, higher LV-MSR was significantly associated with persistent LVH at follow-up (adjusted OR 1.133, 95% CI 1.087–1.180; P<0.001), as well as a higher E/e’ ratio. Associations of LV-MSR at baseline with new-onset and persistent LVH are depicted in **Figure 2**, along with associations of LV-GLS and LV-MI.

**Figure 2.**
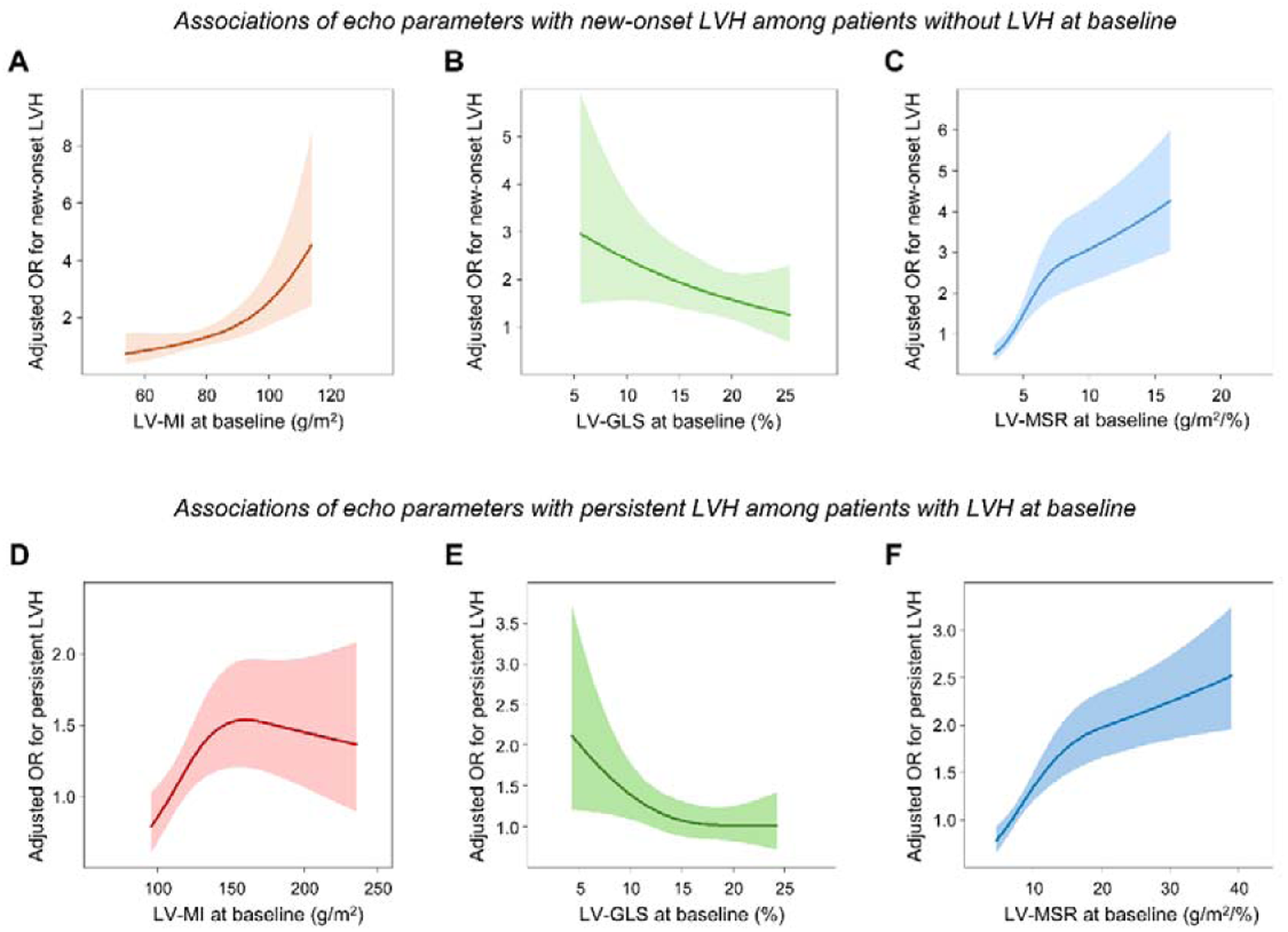
Spline curves of echocardiographic parameters for the changes in LV geometry. Associations of LV-MI **(A)**, LV-GLS **(B)**, and LV-MSR **(C)** at baseline and the new-onset LVH at follow-up echocardiography among the patients without LVH at baseline are shown in the upper panel. Similarly, the associations of LV-MI **(D)**, LV-GLS **(E)**, and LV-MSR **(F)** at baseline and the persistent LVH among those with LVH at baseline are shown in the lower panel. Abbreviations: LVH, left ventricular hypertrophy; OR, odds ratio; others as in Figure 1.

**Table 3.**
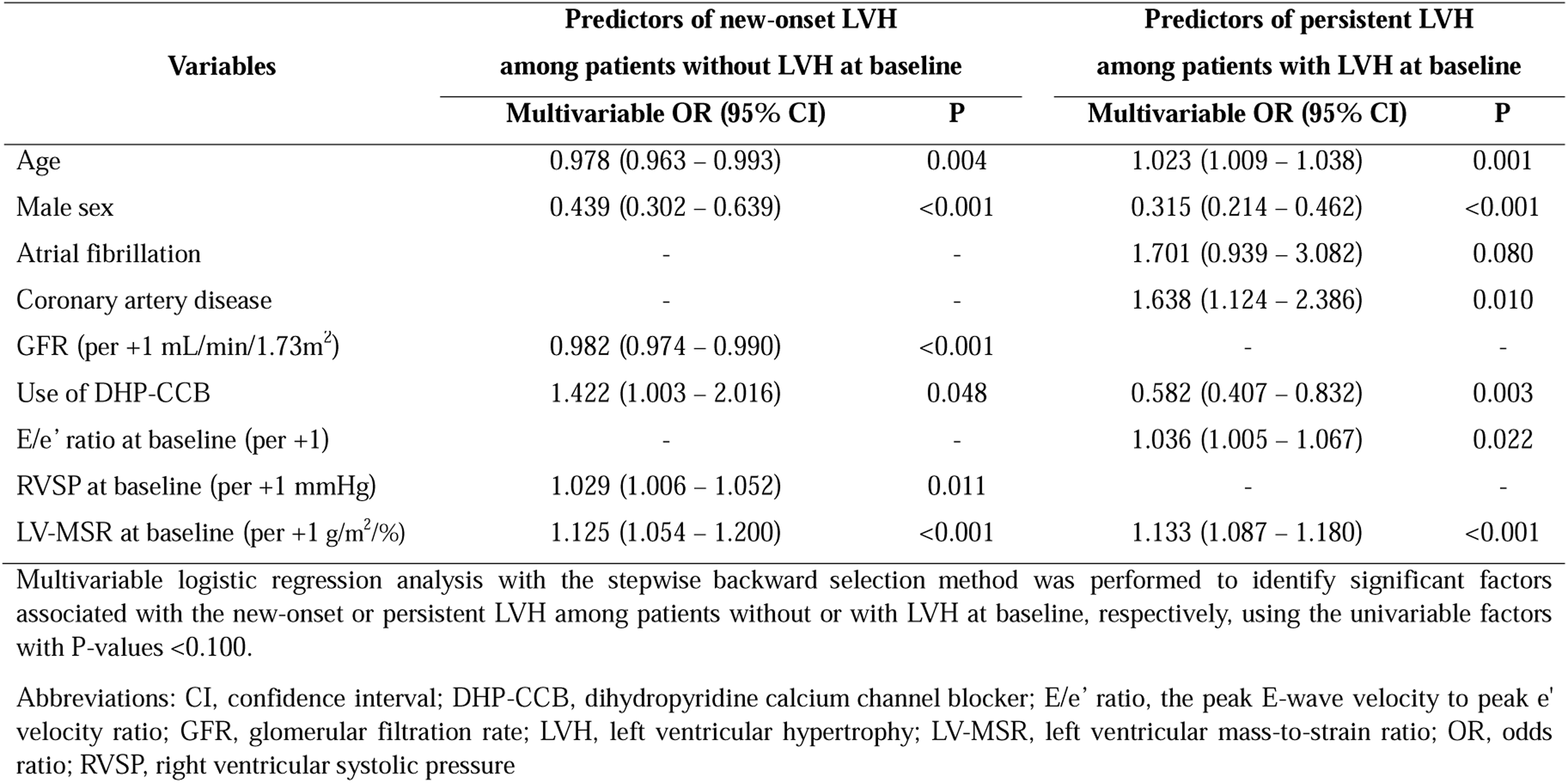
Predictors of new-onset and persistent LVH.

| Variables | Predictors of new-onset LVH<br>among patients without LVH at baseline |  | Predictors of persistent LVH<br>among patients with LVH at baseline |  |
| --- | --- | --- | --- | --- |
|  | Multivariable OR (95% CI) | P | Multivariable OR (95% CI) | P |
| Age | 0.978 (0.963 – 0.993) | 0.004 | 1.023 (1.009 – 1.038) | 0.001 |
| Male sex | 0.439 (0.302 – 0.639) | <0.001 | 0.315 (0.214 – 0.462) | <0.001 |
| Atrial fibrillation | - | - | 1.701 (0.939 – 3.082) | 0.080 |
| Coronary artery disease | - | - | 1.638 (1.124 – 2.386) | 0.010 |
| GFR (per +1 mL/min/1.73m<sup>2</sup>) | 0.982 (0.974 – 0.990) | <0.001 | - | - |
| Use of DHP-CCB | 1.422 (1.003 – 2.016) | 0.048 | 0.582 (0.407 – 0.832) | 0.003 |
| E/e' ratio at baseline (per +1) | - | - | 1.036 (1.005 – 1.067) | 0.022 |
| RVSP at baseline (per +1 mmHg) | 1.029 (1.006 – 1.052) | 0.011 | - | - |
| LV-MSR at baseline (per +1 g/m<sup>2</sup>%) | 1.125 (1.054 – 1.200) | <0.001 | 1.133 (1.087 – 1.180) | <0.001 |

Multivariable logistic regression analysis with the stepwise backward selection method was performed to identify significant factors associated with the new-onset or persistent LVH among patients without or with LVH at baseline, respectively, using the univariable factors with P-values <0.100.
Abbreviations: CI, confidence interval; DHP-CCB, dihydropyridine calcium channel blocker; E/e' ratio, the peak E-wave velocity to peak e' velocity ratio; GFR, glomerular filtration rate; LVH, left ventricular hypertrophy; LV-MSR, left ventricular mass-to-strain ratio; OR, odds ratio; RVSP, right ventricular systolic pressure

### Prognostic value of LV mass-to-strain ratio

ROC curve analysis using bootstrap-based method was performed to assess the prognostic values of the baseline echocardiographic parameters (**Figure 3A**). LV-MSR at baseline revealed a significantly higher AUC for the occurrence of clinical outcomes (AUC 0.696, 95% CI 0.638–0.747), compared to LV-MI (AUC 0.533, 95% CI 0.475–0.590; P-for-comparison <0.001) and LV-GLS (AUC 0.642, 95% CI 0.586–0.697; P-for-comparison <0.001).

**Figure 3.**
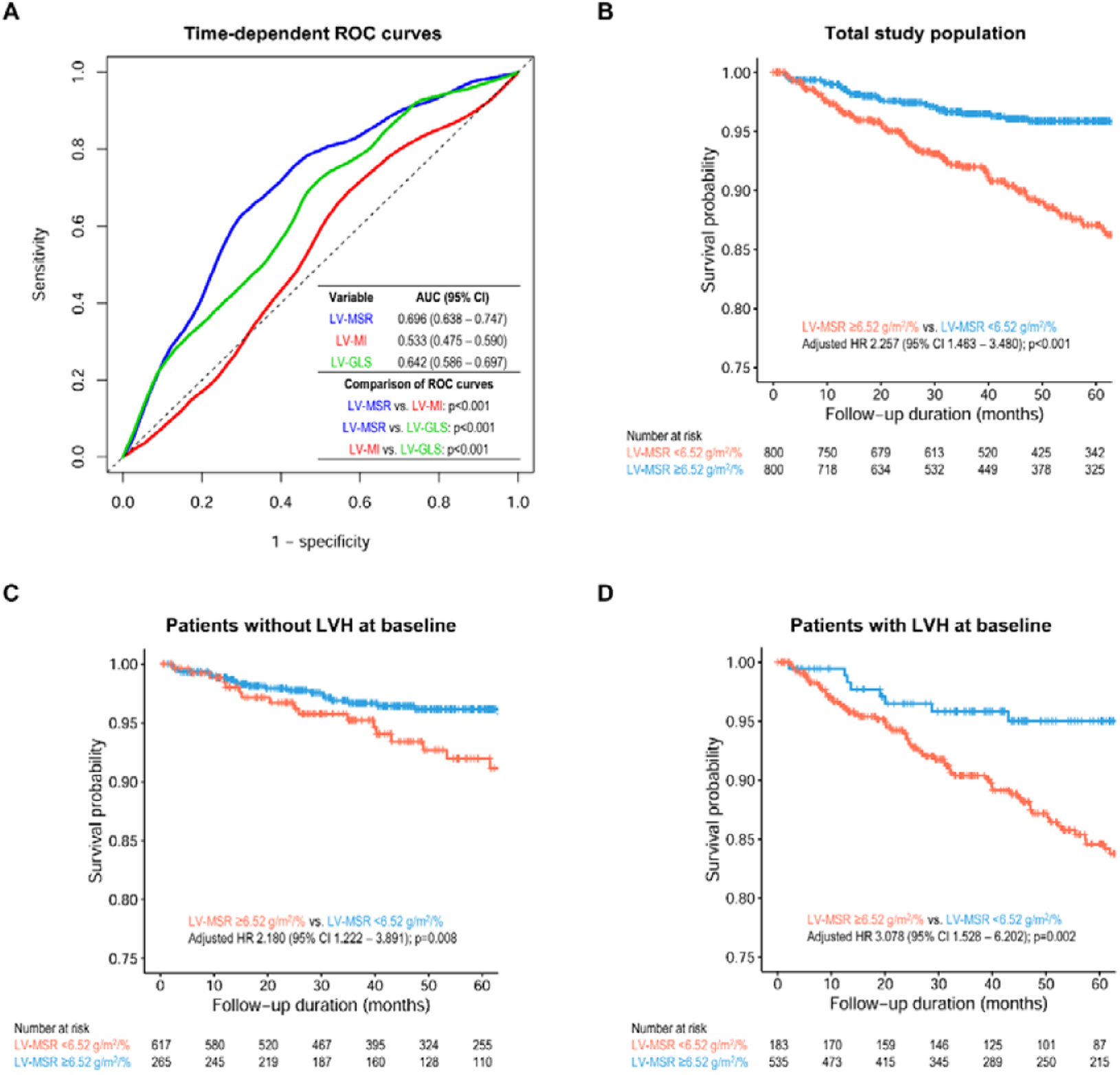
Prognostic value of LV-MSR for cardiovascular death and HHF. **(A)** Time-dependent ROC curves for the occurrence of clinical outcomes (cardiovascular death and hospitalization for heart failure) are compared between LV-MSR, LV-MI, and LV-GLS. The risks of clinical outcomes are compared between high LV-MSR and low-LV-MSR, using the cut-off value of LV-MSR (6.52 g/m^2^/%), among the total study population **(B)**, those without LVH at baseline **(C)**, and those with LVH at baseline **(D)**. Abbreviations: HR, hazards ratio; others as in Figure 1.

Association between LV-MSR at baseline and clinical outcomes was assessed using the multivariable Cox regression analysis (**Table 4** and **Figure 3B**). With LV-MSR as a continuous variable, regardless of the presence of LVH at baseline, higher LV-MSR at baseline was significantly associated with the occurrence of clinical outcomes (adjusted HR 1.032; 95% CI 1.015–1.049; P<0.001) in total study population (without LVH at baseline: adjusted HR 1.048; 95% CI 1.006–1.092; P=0.024 and with LVH at baseline: adjusted HR 1.024; 95% CI 1.005–1.043; P=0.014) (**Table 4**). Using the ROC curve analysis, we determined an optimal cutoff value of LV-MSR for the occurrence of clinical outcomes as 6.52 g/m^2^/%. Higher LV-MSR at baseline (≥6.52 g/m^2^/%) was significantly associated with the occurrence of clinical outcomes (total study population: adjusted HR 2.257; 95% CI 1.463–3.480; P<0.001, without LVH at baseline: adjusted HR 2.180; 95% CI 1.222–3.891; P=0.008, and with LVH at baseline: adjusted HR 3.078; 95% CI 1.528–6.202; P=0.002).

**Table 4.** Predictors of a composite of cardiovascular death and HHF.

| Continuous variables | Total study population |  | Patients without LVH at baseline |  | Patients with LVH at baseline |  |
| --- | --- | --- | --- | --- | --- | --- |
|  | Adjusted HR (95% CI) | P | Adjusted HR (95% CI) | P | Adjusted HR (95% CI) | P |
| Age (per +1 year) | 1.063 (1.040 – 1.086) | <0.001 | 1.060 (1.031 – 1.089) | <0.001 | 1.048 (1.026 – 1.070) | <0.001 |
| BMI (per +1 kg/m<sup>2</sup>) | 1.053 (0.996 – 1.114) | 0.070 | - | - | - | - |
| Diabetes mellitus | 2.093 (1.384 – 3.165) | <0.001 | 1.356 (0.740 – 2.487) | 0.324 | - | - |
| Chronic kidney disease | 1.768 (1.144 – 2.731) | 0.010 | - | - | 1.895 (1.209 – 2.971) | 0.005 |
| Coronary artery disease | 1.606 (1.056 – 2.443) | 0.027 | 1.368 (0.755 – 2.477) | 0.301 | - | - |
| Use of MRA | - | - | 2.367 (0.725 – 7.726) | 0.154 | - | - |
| E/e' ratio at baseline (per +1) | 1.031 (1.011 – 1.052) | 0.002 | - | - | - | - |
| RVSP (per +1 mmHg) | - | - | - | - | 1.023 (1.007 – 1.038) | 0.003 |
| LV-MSR at baseline (per +1 g/m<sup>2</sup>/%) | 1.032 (1.015 – 1.049) | <0.001 | 1.048 (1.006 – 1.092) | 0.024 | 1.024 (1.005 – 1.043) | 0.014 |
| Categorical variables | Total study population |  | Patients without LVH at baseline |  | Patients with LVH at baseline |  |
|  | Adjusted HR (95% CI) | P | Adjusted HR (95% CI) | P | Adjusted HR (95% CI) | P |
| Age ≥70 years | 2.341 (1.635 – 3.354) | <0.001 | 2.655 (1.450 – 4.861) | 0.002 | 2.471 (1.578 – 3.868) | <0.001 |
| Diabetes mellitus | 1.988 (1.305 – 3.028) | 0.001 | 1.388 (0.766 – 2.515) | 0.280 | - | - |
| Chronic kidney disease | 1.745 (1.140 – 2.670) | 0.010 | - | - | 2.010 (1.300 – 3.107) | 0.002 |
| LAVI ≥40 mL/m<sup>2</sup> | 1.875 (1.160 – 3.030) | 0.010 | 1.618 (0.858 – 3.052) | 0.137 | - | - |
| RVSP ≥40 mmHg | 1.754 (1.081 – 2.847) | 0.023 | - | - | 2.633 (1.667 – 4.158) | <0.001 |
| LV-MSR ≥6.52 g/m<sup>2</sup>/% | 2.257 (1.463 – 3.480) | <0.001 | 2.180 (1.222 – 3.891) | 0.008 | 3.078 (1.528 – 6.202) | 0.002 |

Multivariable Cox proportional hazards regression models were performed using the stepwise backward selection method, including variables that were significant in the univariable analysis. The upper section of the table presents results with all variables, including LV-MSR, analyzed in their continuous form, while the lower section displays results with all variables analyzed in their categorical form
Abbreviations: adjusted HR, adjusted hazard ratio; BMI, body mass index; CI, confidence interval; E/e' ratio, the peak E-wave velocity to peak e' velocity ratio; HHF, hospitalization for heart failure; LVH, left ventricular hypertrophy; LV-MSR, left ventricular mass-to-strain ratio; MRA, mineralocorticoid antagonist; RVSP, right ventricular systolic pressure

Because higher LV-MSR was significantly associated with new-onset or persistent LVH at follow-up echocardiogram (which are considered as prognostic markers for clinical outcomes), we performed mediation analysis to assess the direct and indirect effects of LV-MSR through its influence on LV geometry changes (**Figure 4**). The mediation analysis showed that 47% of the effect of LV-MSR at baseline on clinical outcomes was mediated by the changes in LV geometry (new-onset or persistent LVH at follow-up echocardiogram).

**Figure 4.**
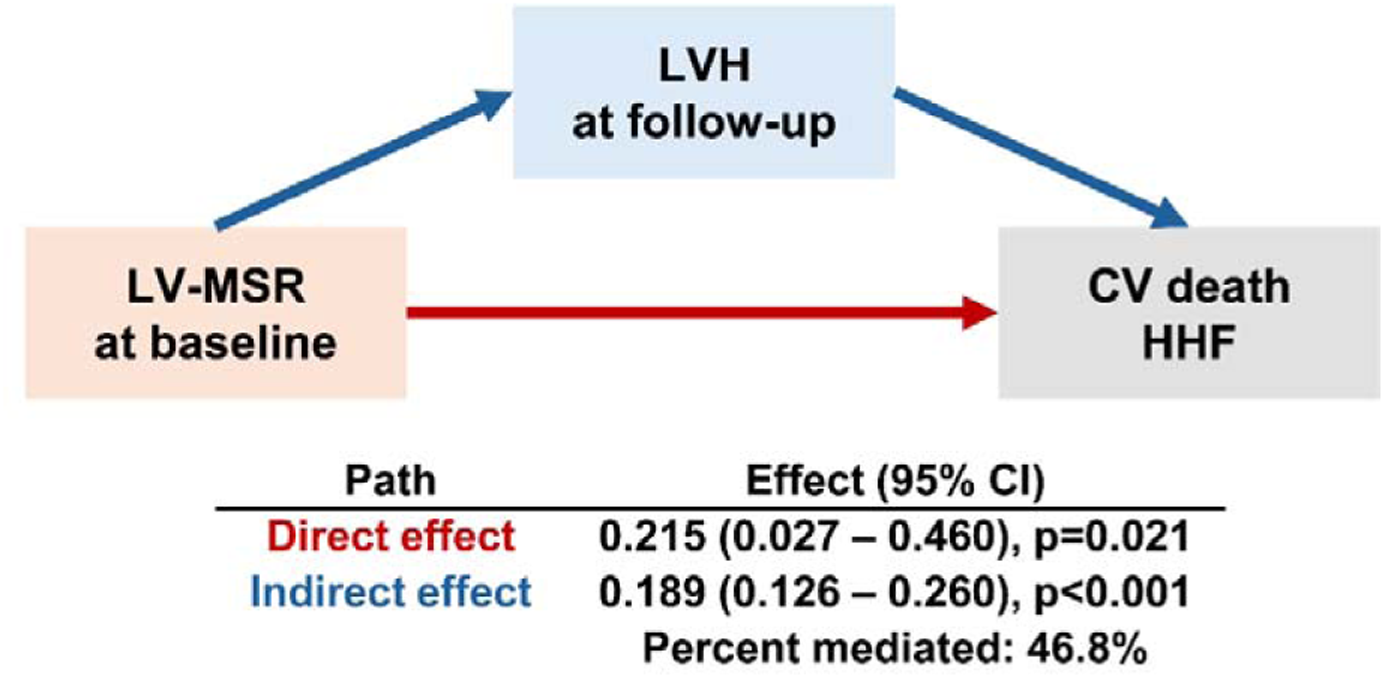
Mediation analysis on the association between LV-MSR, changes in LV geometry, and prognosis. Abbreviations: ACME, average causal mediation effect; ADE, average direct effect; CV, cardiovascular; HHF, hospitalization for heart failure; LVH, left ventricular hypertrophy; LV-MSR, left ventricular mass-to-strain ratio

## Discussion

In the present study, we assessed the impacts of LV-MSR, the ratio between LV-MI and LV-GLS, on the changes in LV geometry and prognosis among the patients diagnosed with hypertension. The higher LV-MSR (i.e., disproportionately higher LV-MI compared to LV-GLS) was significantly associated with new-onset LVH among those without LVH at baseline, and with persistence of LVH among those with LVH at baseline. Further, LV-MSR had a higher predictive value for the changes in LV geometry, compared to LV-MI and LV-GLS when used separately. Higher LV-MSR was also significantly associated with the occurrence of clinical outcomes, mediated partly by changes in LV geometry. To our knowledge, this study is the first to demonstrate the clinical usefulness of LV-MSR among patients with hypertension, showing that LV-MSR is both a predictor of the changes in LV geometry and a prognostic marker of clinical events. Our findings suggest that LV-MSR could be used as a relevant marker to assess treatment response and estimate prognosis in patients with hypertension.

### Echocardiographic assessment in patients with hypertension

According to previous studies, the prevalence of LVH ranges from 15% to 45% among patients with hypertension. Given that LVH is a physiologic adaptation to increased afterload in these patients, the presence of LVH serves as a key component to determine HHD, as well as an important marker of prognosis.^1,2^ Moreover, our previous studies showed that appropriate BP control could result in the regression of LVH, thus leading to improved prognosis, whereas failure to achieve LVH regression results to a worse prognosis.^10,11^

While most of previous literature focused on changes in LV geometry and LVH status, we extended the clinical implications of HHD using LV strain measurements. The well-established better sensitivity of LV-GLS measurement for assessing subclinical myocardial dysfunction, over conventional LV-EF, has also been confirmed in patients with HHD. A meta-analysis of 4,276 individuals (2,187 patients with hypertension) reported similar LV-EF between normotensive and hypertensive patients, although LV-GLS was significantly impaired in the hypertensive group, with a standard mean difference of 1.07±0.15 (95% CI 0.77–1.36, P<0.0001).^7^ Further, an in-depth mathematical analysis of the associations between LV geometry, LV-EF, and strain measurements by Stokke et al. showed that LV-EF can be maintained in a setting of increased LV wall thickness despite a significant impairment of longitudinal deformation (i.e., LV-GLS).^15^ This implies that LV-GLS may be a more sensitive marker of LV dysfunction in patients with HHD. Although further clarification is needed for a wider utilization of LV-GLS in patients with HHD, contemporary literature suggests that LV-GLS has potential of clinical relevance in the assessment of HHD.^2-4^

While we recognize the potential of LV-GLS in the assessment of HHD, an important feature of strain measurements in a setting of hypertrophied myocardium exists. Overall, it could be summarized that the adaptive LV remodeling to increased afterload results in the simultaneous increase in LV wall thickness (i.e., LVH) and impairment of longitudinal function (i.e., LV-GLS). This was demonstrated in a landmark mechanistic analysis of LV geometry and function in patients with HHD by Hare et al., which showed a step-wise decrease in LV-GLS in patients with concentric remodeling and concentric hypertrophy compared to those with normal geometry.^5^ However, it should be noted that, while the LV longitudinal function decreases as HHD progresses, the overlap in LV-GLS values between the phenotypic (or geometric) subgroups is substantial. Similar findings were also reported by Le et al., showing that an impaired LV-GLS in hypertensive patients with LVH, still shows significant overlap between subgroups.^16^ Tadic et al. also showed significant but modest correlations of LV-MI with LV-GLS in patients with hypertension with coefficients ranging from -0.39 to -0.44.^17^ In addition, among healthy individuals enrolled to determine the reference values of LV strain, there was only a weak correlation between LV mass and LV-GLS, which was finally nullified when LV-MI, instead of LV mass, was used.^18^ These findings suggest that, while the physiologic adaptation to increased afterload in patients with HHD involve both the hypertrophy of myocardium and impairment of longitudinal function, these two pathologic processes are not always proportional. In other words, the stages of HHD and its consequences cannot be captured by the single use of either LV-MI or LV-GLS, but could be better determined using both parameters.

In the present study, we found a significant but weak correlation between LV-MI and LV-GLS (Pearson’s correlation coefficient r = -0.345, P<0.001) (**Supplemental Figure S1**). Further, while both LV-GLS and LV-MI were significantly associated with the presence of LVH at follow-up echocardiogram, LV-MSR revealed a significantly higher AUC value compared to either LV-GLS or LV-MI, and showed independent predictive value for new-onset LVH among those without LVH at baseline, as well as for persistent LVH among those with LVH at baseline. Our findings indicate that the combination of both LV-MI and LV-GLS, in the form of their ratio (LV-MI/LV-GLS; LV-MSR), could better reflect the myocardial status in patients with HHD, providing relevant prediction of changes in LV geometry after anti-hypertensive treatment.

Furthermore, several recent studies suggest the use of LV-MSR in differential diagnosis of thick myocardium, including the differentiation of light chain (AL) versus transthyretin (ATTR) cardiac amyloidosis and Fabry disease from HHD.^8,9^ According to Ferkh et al., LV-MSR values were significantly lower in patients with HHD (median 4.95, interquartile range [IQR] 4.31–5.56) compared to those with cardiac amyloidosis or Fabry disease.^8^ In the present study, the mean LV-MSR at baseline was significantly higher (mean 8.79, SD 7.07) than that by Ferkh et al., and LV-MI was also higher (mean 109.8, SD 33.7) than Ferkh et al.’s (median 92.7, IQR 79.6–102.3); however, LV-GLS was lower (mean 15.3, SD 5.0 vs. median 18.6, IQR 17.5–20.0, respectively). These findings suggest that the patients included in our study had more advanced phenotypes of HHD, considering that our study included those with hypertension referred to tertiary institutions and underwent repetitive echocardiograms.

### Hypertensive heart disease: prognostic value of LV-MSR

According to Saito et al., LV-GLS and presence of concentric hypertrophy had significant prognostic value for the occurrence of major adverse events among asymptomatic patients with HHD.^4^ In particular, in the “Echo Model” based on multivariable Cox regression analysis, the presence of concentric hypertrophy had an adjusted HR of 1.75 (95% CI 1.08–2.84; P=0.02) while for LV-GLS, it was 1.08 (95% CI 1.01–1.17; P=0.04) per 1% decrease in value. This finding is noteworthy in that both LVH and LV-GLS revealed independent prognostic values, highlighting the importance of assessing both the structural change and functional impairment in patients with HHD.

Adding to this previous finding, we demonstrated that the ratio of LV-MI with LV-GLS (LV-MSR) has a significant prognostic value for the occurrence of CV death and HHD, regardless of whether analyzed as a continuous or categorical variable. In addition, we conducted mediation analyses to investigate whether LV-MSR is directly associated with prognosis, or if it primarily predicts changes in LV geometry, indirectly influencing clinical outcomes. The results indicated that LV-MSR at baseline had both a direct prognostic effect and an indirect effect by predicting LV geometry at follow-up echocardiography. These findings may explain why LV-MSR outperforms LV-MI or LV-GLS alone in prognostic assessment. Moreover, they suggest that the incorporation of LV function (LV-GLS) into LV geometry (LVH) could serve as both a predictor of treatment response and a prognostic marker in patients with HHD.

### Limitations

The present study has several limitations. First, because the study population was derived from tertiary referral hospitals, there may have been a selection bias between our study population and general patients with hypertension. Secondly, the exact duration of hypertension could not be confirmed because of the retrospective study design. Third, we could not quantitatively measure irreversible myocardial fibrosis because we utilized echocardiography rather than cardiac magnetic resonance (CMR) imaging. Finally, given the retrospective design of this study, the causal relationship between LV-MSR and the changes in LV geometry, as well as the occurrence of clinical events, cannot be confirmed, requiring future prospective studies.

### Conclusions

In patients with hypertension, the ratio between LV-MI and LV-GLS (i.e., LV-MSR) provides independent predictive value for changes in LV geometry, and can be used as a relevant prognostic marker following antihypertensive treatment. The use of LV-MSR, in comparison to using either LV-MI or LV-GLS, could better reflect the treatment response and clinical course in patients with HHD.

## Data Availability

All data produced in the present study are available upon reasonable request to the authors.

## Acknowledgments

We thank Lia Ju, a registered diagnostic cardiac sonographer (RDCS), and Eun-Ju Choi, a research nurse, for their dedication and support.

## Abbreviations

ACME: average causal mediation effect
ADE: average direct effect
AUC: area under the curve
BMI: body mass index
CI: confidence interval
HHF: hospitalization for heart failure
LAVI: left atrial volume index
LV: left ventricle
LVH: left ventricular hypertrophy
LV-MI: left ventricular mass index
LV-GLS: left ventricular global longitudinal strain
LV-MSR: left ventricular mass-to-strain ratio
ROC: receiver-operating characteristics
RWT: relative wall thickness

