## Supplemental Table S1 and Supplemental Figure S1 for "Left ventricular mass-to-strain ratio to predict treatment response and prognosis in hypertensive heart disease"

**Short Title:** LV mass-to-strain ratio predicts outcomes of HHD

In-Chang Hwang^a,b*^, Hyue Mee Kim^c*^, Jiesuck Park^a^, Hong-Mi Choi^a,b^, Yeonyee E. Yoon^a,b^, Goo-Yeong Cho^a,b^

^a^ Department of Cardiology, Cardiovascular Center, Seoul National University Bundang Hospital, Seongnam, Gyeonggi;

^b^ Department of Internal Medicine, Seoul National University College of Medicine, Seoul;

^c^ Division of Cardiology, Department of Internal Medicine, Chung-Ang University Hospital, Seoul, South Korea

^*^ These two authors equally contributed to this work as co-corresponding authors**.**

**Supplemental Table S1. Multiple linear regression analysis for prediction of the reduction in LV-MI (△LV-MI)**

| **Variables** | **Coefficient β** | **95% CI** | **p** |
| --- | --- | --- | --- |
| Age (per +1 year) | 0.120 | -0.020 – 0.261 | 0.094 |
| BMI (per +1 kg/m^2^) | 0.554 | 0.087 – 1.020 | 0.020 |
| SBP (per +1 mmHg) | -0.139 | -0.211 – -0.067 | <0.001 |
| Diabetes mellitus | 2.057 | -1.491 – 5.605 | 0.256 |
| Chronic kidney disease | 3.194 | -0.739 – 7.128 | 0.111 |
| Use of beta blockers | -1.931 | -5.268 – 1.407 | 0.257 |
| Use of DHP-CCB | -5.375 | -8.687 – -2.063 | 0.001 |
| LV-EDV (per +1 mL) | -0.204 | -0.262 – -0.145 | <0.001 |
| RWT (per +1) | -60.589 | -77.243 – -43.935 | <0.001 |
| LV-MSR (per +1 g/m^2^/%) | -0.602 | -0.875 – -0.330 | <0.001 |

Abbreviations: BMI, body mass index; CI, confidence interval; LV-EDV, left ventricular end-diastolic volume; LV-MI, left ventricular mass index; LV-MSR, left ventricular mass-to-strain ratio; RWT, relative wall thickness; SBP, systolic blood pressure

**Supplemental Figure S1. Correlation between LV-MI and LV-GLS**

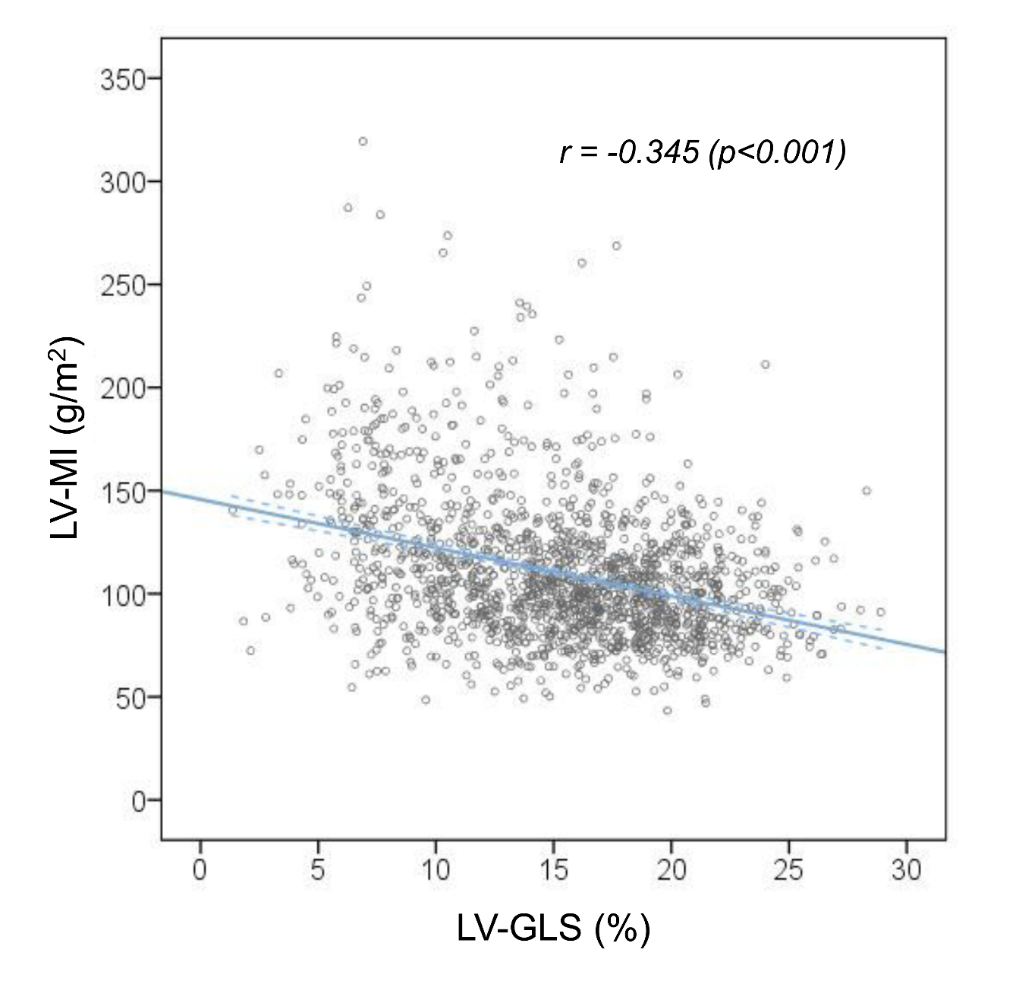

Abbreviations: LV-MI, left ventricular mass index; LV-MSR, left ventricular mass-to-strain ratio
